# Prognostic value of the preoperative systemic immune-inflammation index for overall survival after Surgical resection in gallbladder cancer: a systematic review and meta-analysis

**DOI:** 10.64898/2025.12.16.25342369

**Authors:** Mohammadsadra Shamohammadi, Armaghan Abbasi Garavand, Seyedeh Mohadese Mosavi Mirkalaie, Amirarsalan Varmahziar, Mansour Bahrdoust

## Abstract

**Background:** The systemic immune-inflammation index (SII) reflects the relationship between tumor-promoting inflammation and anti-tumor immunity in various solid malignancies, but the role of SII in Gallbladder cancer (GBC) has yet to be established. The aim of the systematic review and meta-analysis was to clarify the prognostic value of preoperative SII in GBC patients undergoing resection.

**Methods:** This systematic review and meta-analysis was performed following PRISMA 2020 checklist and we searched PubMed, Embase, Web of Science and Scopus from inception to November 1, 2025. Original studies with English language enrolled adults with resectable GBC undergoing surgical resection and reported preoperative SII with overall survival (OS) as a hazard ratio (HR) comparing high versus low SII. Risk of bias was assessed with the Newcastle-Ottawa Scale (NOS). Random-effects models were applied to pool HRs, and heterogeneity was summarized with I² and τ². We evaluated publication bias with visual inspection of funnel plots and Egger’s regression test.

**Results:** Seven studies (N= 2,153) met inclusion criteria. High preoperative SII was associated with significantly worse OS (pooled HR 2.17; 95% CI 1.55–2.79). with moderate heterogeneity (I² = 26.0%, τ² = 0.0285). Results were robust in leave-one-out analyses, and variability in study-specific SII cut-offs accounted for part of the heterogeneity. Certainty of evidence for the primary outcome was moderate, and all included studies were high quality.

**Conclusions:** preoperative SII is an inexpensive, available biomarker that correlated with risk in resectable GBC and is able to identify patients with more aggressive tumor biology despite Surgical surgery.

**Systematic review registration:** PROSPERO CRD420251185808

## Background

Gallbladder cancer (GBC) is a relatively rare type of cancer, which this type of malignancy causes about 1.7% of cancer mortality worldwide in 2018 (1, 2). The aggressive biological behavior often leads to the late and clinically significant diagnosis of the condition, thus being associated with a poor prognosis (1, 3). Despite current therapeutic interventions, postoperative recurrence rates and overall survival (OS) outcomes remain unsatisfactory (4). Therefore, careful disease assessment and the identification of novel prognostic biomarkers are necessary to achieve optimized therapeutic targeting and patient outcomes in GBC (5, 6).

In this context, chronic inflammation is recognized as one of the key contributors to gallbladder carcinogenesis and tumor progression because inflammation-inducing responses to lithogenic bile, gallstones, environmental toxin exposure, and infectious or autoimmune disorders can trigger epithelial remodeling, which progresses into invasive carcinoma (7, 8). In addition, inflammation facilitates malignant transformation through processes that include the induction of genetic mutations, clonal growth of malignant cells, angiogenesis, and the controlled spread of metastases (9). Consequently, several inflammation-related indices and hematological biomarkers, including the neutrophil-lymphocyte ratio, platelet distribution width, fibrinogen-to-albumin ratio, and preoperative platelet count, have progressively greater attention as prognostic factors in GBC (10–12).

The systemic immune-inflammation index (SII), which is calculated from platelet, neutrophil, and lymphocyte count, has been developed as a valuable biomarker that reflects both the host immune status and systemic inflammatory response (13, 14). Recent evidence has shown higher SII associated with poor prognosis outcomes across various malignancies, including gastrointestinal, ovarian, breast, and lung cancer (13, 15–19). However, the hazard ratio of higher SII on survival rate in GBC has been heterogeneous and requires clarification (20–23), which may be low power of the individual studies. Therefore, to address this gap, we conducted a systematic review and meta-analysis to clarify the prognostic value of preoperative SII for OS in GBC patients undergoing surgical resection.

## Methods

### Literature search

This systematic review and meta-analysis followed the Preferred Reporting Items for Systematic reviews and Meta-Analyses (PRISMA) 2020 reporting guideline (24), and the protocol was prospectively registered in PROSPERO (CRD420251185808). We conducted comprehensive searches in PubMed, Embase, Web of Science, and Scopus databases from their inception through November 1, 2025. Based on the study PICO framework, the database search strategies combined controlled vocabulary and keywords related to “gallbladder cancer” and “systemic immune-inflammation index”. Full search terms for each database are detailed in Supplementary Table S1. No restrictions on study design and language publication were applied at the search stage. Reference lists of included studies and relevant reviews were hand-searched. Complete search strategies are provided in Supplementary Table 1.

### Eligibility criteria

The inclusion parameters used to select eligible studies were as follows: (1) studies with adult patients (≥18 years) with histopathologically confirmed resectable GBC who underwent surgical resection, (2) availability of preoperative SII values (calculated as platelet count × neutrophil count / lymphocyte count), (3) reporting of overall survival outcome in relation to SII, such as a hazard ratio (HR) comparing high versus low SII, and (4) original research articles with an observational cohort or randomized trial design with complete clinical and follow-up data.

We excluded studies that: (1) involved non-resection populations, (2) lacked preoperative SII data or relevant survival outcomes, (3) did not provide prognostic outcomes, (4) contained duplicated or overlapping data, or (5) were not published in English, or were abstracts, letters, editorials, reviews, case reports, or case series.

### Study selection and data extraction

Two independent reviewers screened the titles and abstracts of all the identified studies after removing duplicate records. After that, Full texts of potentially relevant studies were reviewed by a third reviewer based on predefined inclusion and exclusion criteria, with disagreements resolved by a fourth reviewer. In the selected studies, extensive data extraction was done, documenting all important variables, such as study characteristics (first author, publication year, country), design and setting, enrollment period, sample size, follow-up duration; patient demographics (age, sex), histology, stage, treatment modalities; SII cut-off values; outcomes including hazard ratios (HR) with 95% confidence intervals (CI), and 5-year OS and recurrence-free survival (RFS) stratified by SII; and effect sizes. Data were independently verified by two researchers for accuracy and consistency, with discrepancies resolved through consultation with a third team member.

### Quality assessment

The methodological quality of the cohort studies included was used to determine the Newcastle-Ottawa Scale (NOS), a validated tool that evaluates the selection of participants, the comparability of cohorts, and the ability to measure the outcomes, with a maximum score of nine (25). The appraisal of each study was done in terms of design, sample size, data collection method, and confounding factors. NOS scores were independently rated by two reviewers as high quality (7–9), moderate quality (4–6), or low quality (0–3). The revised Cochrane Risk of Bias 2 (RoB 2) tool was used to determine the quality of the randomized controlled trials (26). It is a tool that analyses bias in five areas, including selection of reported results, no outcome data, not as intended, randomization process introduces bias, and outcome measurement. Bias levels were determined based on structured signaling questions as low, some concerns, and high per domain. Each assessment was conducted by two reviewers, and any disagreements were to be resolved by discussing them or using the services of a third reviewer. In this review, all studies that scored above four were included the study.

### Certainty of evidence

The confidence of the evidence was also evaluated with the help of the GRADE tool (27). It focuses on multiple critical domains such as risk of bias, inconsistency, indirectness, imprecision, and publication bias. To indicate the level of evidence on each outcome, the evaluation of its overall quality grade is provided in four levels: high, moderate, low, and very low. The included observational studies were subsequently reassessed for possible upgrading or downgrading according to the domains. This approach to methodology made a very rigid and transparent assessment of effect estimates possible, thus influencing the interpretation of the findings of the meta-analysis and the strength of the resulting conclusions.

### Statistical analyses

Data were analyzed with Stata version 17. We analyzed the data using a random model to control for the effect of sample size of included studies on the overall data estimates. The effect size of interest was the hazard ratio (HR) for high SII versus low SII. We reported the pooled HR with 95% confidence interval. Statistical heterogeneity was assessed using the I² statistic (with values >40% indicating significant heterogeneity) and the between-study variance τ². We used sensitivity analysis to assess the effects of individual studies. Potential publication bias was assessed with the Egger test and the results are presented in a funnel plot. Given the absence of publication bias, trim and fill analyses were not required. Given the low level of heterogeneity between studies and the limited number of studies (<10 studies), we did not perform meta-regression analyses.

## Results

### Study selection

We identified 206 records from databases (PubMed n=30, Embase n=51, Scopus n=93, Web of Science n=32). After removing 87 duplicates, 119 records were screened and 98 were excluded. We sought and retrieved 21 reports for full-text review, assessed 21 full texts for eligibility, and excluded 14. Seven studies met inclusion criteria and were included in the meta-analysis (Figure 1).

**Figure 1:**
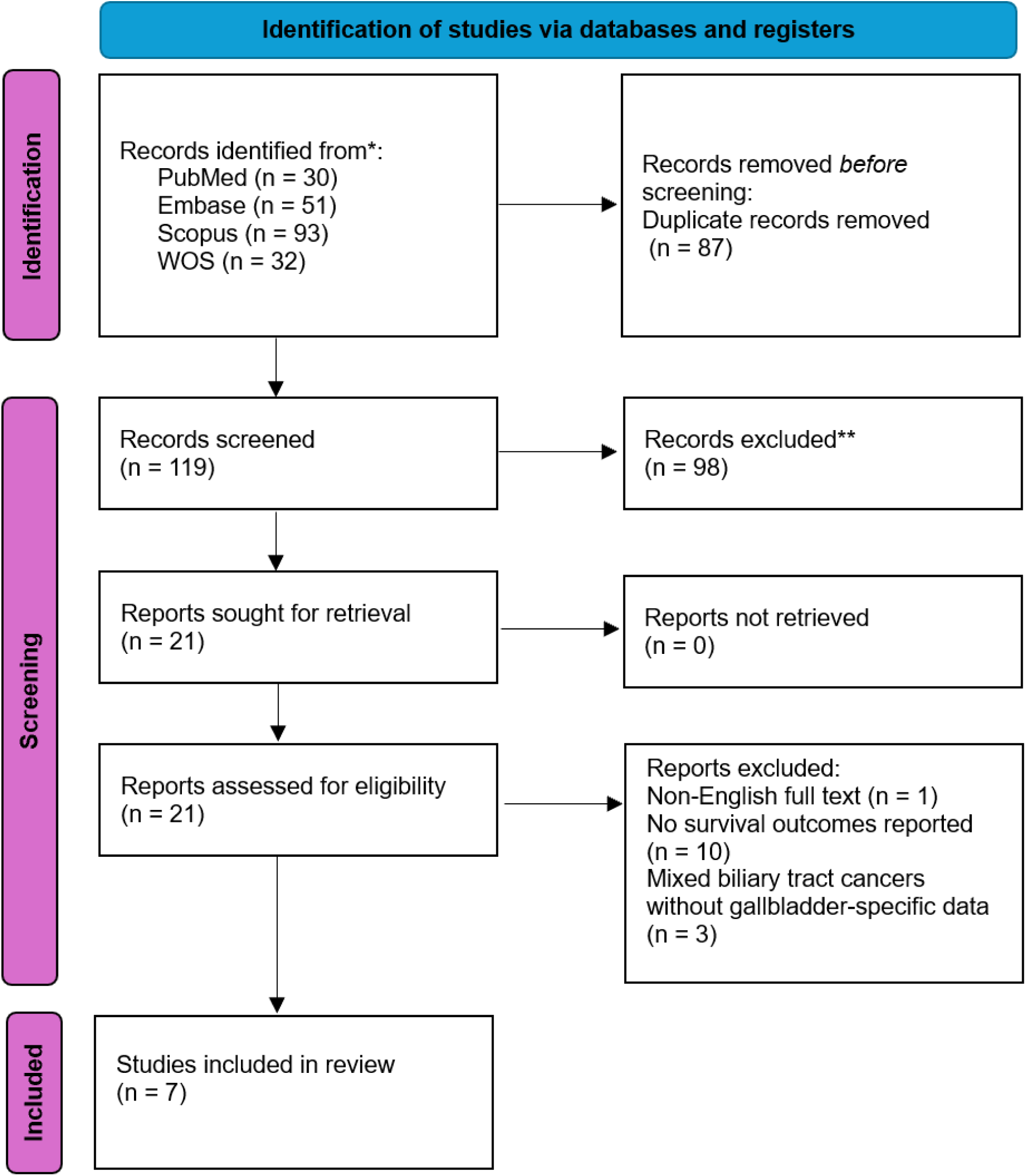
PRISMA 2020 flow diagram of study selection for the meta-analysis.

### Study characteristics

Seven cohorts (11, 20–23, 28, 29) (n = 2,153) from China (publication years 2020–2023) were included (Table 1). Across studies that reported sex (n = 2,027 patients), 39.2% were male (794/2,027) and 60.8% female (1,233/2,027). 72.0% of patients had TNM stage III–IV (1,188/1,652), and where nodal status was reported (n = 1,458), 35.6% were lymph-node positive (519/1,458). All studies evaluated preoperative SII as a dichotomous exposure. The median cut-off was 600 with IQR (510–829.2).

**Table 1:** Baseline characteristics of included cohorts.

| Author (year) | Type of study | Country | Period | No total | No high/low cut-off | Gender (male/female) | Age (Mean±SD) | TNM stage I-II/III-IV | Lymph node (+/-) | Perineural invasion (yes/no) | R0 | Cutoff-selection | Cut-off |
| --- | --- | --- | --- | --- | --- | --- | --- | --- | --- | --- | --- | --- | --- |
| Huang (2025)(20) | Multicenter Retrospective cohort | China | 2015-2023 | 210 | 121/89 | 80/130 | 60.75±15 | 109/101 | 64/146 | 9/125 | NA | ROC | 590.105 |
| Li (2023)(11) | Single-center Retrospective cohort | China | 2011-2020 | 427 | 190/110 | 92/208 | NA | 57/243 | 75/225 | NA/NA | NA | ROC | 399.7 |
| Liu (2025)(21) | Single-center Retrospective cohort | China | 2015-2017 | 86 | 40/46 | 33/53 | NA | 33/53 | 39/47 | 30/56 | 86 | ROC | 889.52 |
| Sun (2020)(29) | Single-center Retrospective cohort | China | 2003-2017 | 142 | 70/72 | 60/82 | 63.06±10.68 | 24/113 | 64/78 | NA/NA | 88 | Harrell's C and Somers' D statistical tests | 600 |
| Yin (2024)(22) | Multicenter Retrospective cohort | China | 2013-2022 | 123 | 34/89 | 53/70 | 65.7±9.6 | 43/80 | 49/74 | 35/88 | 115 | ROC | 829.2 |
| Li (2021)(28) | Multicenter Retrospective cohort | China | 2002-2019 | 1072 | 550/522 | 414/658 | 63.0±9.4 | 158/617 | 200/304 | 118/519 | 65 | Cut p function (R package survMisc) | 510 |
| Chen (2021)(23) | Single-center Retrospective cohort | China | 2012-2020 | 93 | 43/50 | 62/31 | 62.0±14.5 | 40/53 | 28/65 | NA/NA | NA | ROC | 823.99 |

### Risk of bias

Using the NOS, overall study quality was high in most cohorts (≥7 stars), with common limitations in selection and adjustment domains (retrospective design; incomplete covariate control). Full NOS ratings by domain are provided in Table S2.

### Association between SII and overall survival

Pooled results showed that high preoperative SII versus low SII was significantly associated with worse OS in gallbladder cancer (HR = 2.17 (95% CI 1.55-2.79) (Figure 2). Heterogeneity between studies was acceptable and small (I² = 26.0).

**Figure 2:**
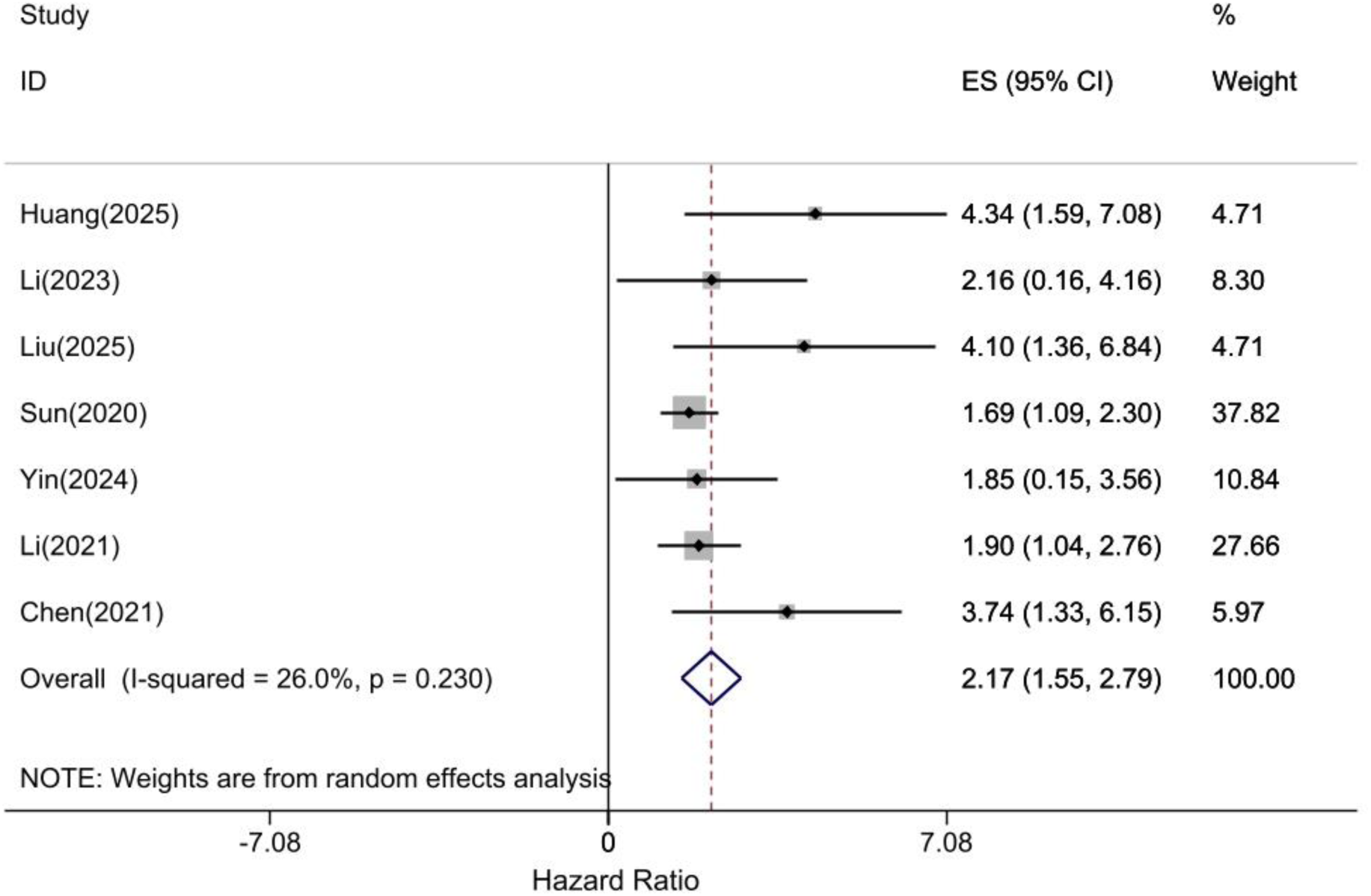
Forest plot of overall survival comparing high vs low preoperative SII after *Surgical* resection; individual and pooled HRs (95% CI) shown (random-effects model).

### Sensitivity analysis

Sensitivity analysis was performed to assess the impact of individual studies on the outcome of the association of SII with patient survival (Figure S1). The results of the sensitivity analysis showed that studies with a cut-off value ≤600 had the greatest impact on the overall outcome of the association between SSI and OS.

### Publication bias

Visual inspection of the funnel plot (Figure 3) did not reveal marked asymmetry. Egger’s test showed no evidence of small-study effects (t = 1.5, 95% CI −0.1 to 3.1, P = 0.094).

**Figure 3:**
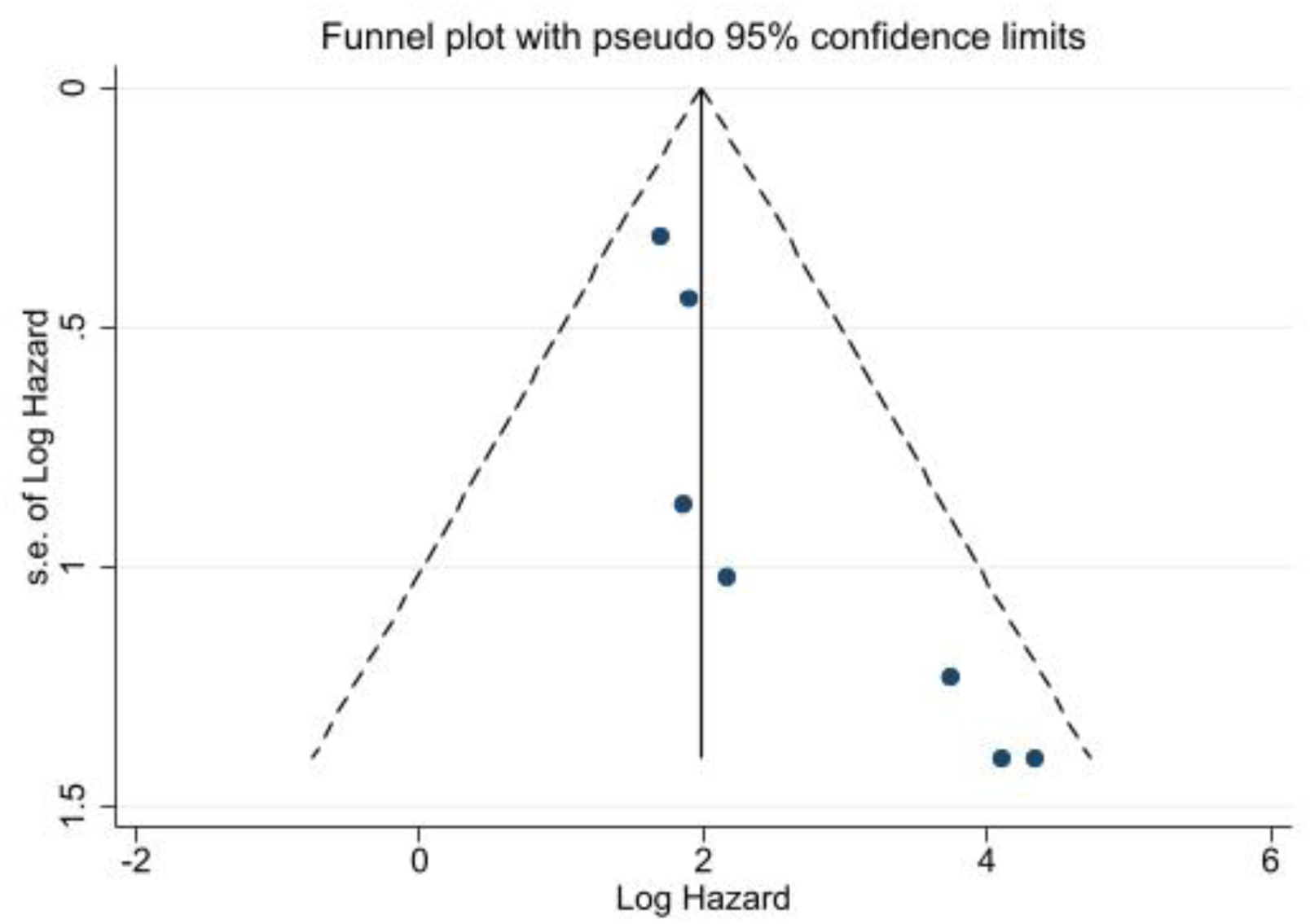
Funnel plot of included studies (log [HR] vs SE) assessing small-study effects; no marked asymmetry observed.

### Certainty of evidence (GRADE)

Applying GRADE for prognosis, certainty in the association between higher preoperative SII and shorter overall survival was rated moderate, downgraded one level for risk of bias (retrospective design and limited adjustment in several studies). No downgrades were applied for inconsistency, indirectness, or imprecision. The Summary of Findings is shown in Table S2.

## Discussion

The present systematic review and meta-analysis demonstrates that a high preoperative SII is associated with significantly worse OS following Surgical resection of GBC. The association was robust across sensitivity analyses and was observed moderate heterogeneity, which was attributable to variability in study-specific SII cut-offs. Study quality was consistently high (NOS ≥7), and the certainty of evidence for the primary outcome was moderate according to GRADE. This suggests that patients with high SII have over two times the risk of poor OS than patients with low SII. The effect size quantifies the clinical significance of systemic inflammation to survival outcomes in the resectable setting, and corroborate findings of individual studies. In a retrospective analysis of patients undergoing Surgical resection for GBC, Li et al. (30) evaluated SII levels ≥510 and found a significant association for shorter median OS, nearly 90% higher mortality hazard on multivariate analysis. Similarly, Chen et al. (31) demonstrated that elevated SII associated with worse long-term outcomes following surgical resection. Our meta-analysis synthesizes evidence that the SII is a prognostic biomarker in resectable GBC, and that elevated preoperative SII may help identify patients with more aggressive tumor biology and poorer overall survival

Established markers including the NLR, PLR, and MLR have been shown to be prognostic for survival in GBC, but the SII includes information of both NLR and PLR and may have greater prognostic value (32–34). In a previous meta-analysis of published studies of hematologic indices in GBC the pooled HR for poor OS for elevated NLR was 1.72 (95% CI, 1.47–2.02), for elevated PLR 1.51 (95%, CI, 1.33–1.72), and for elevated MLR was 1.96 (95% CI, 1.46–1.64) (32). In our analysis, the HR for SII was larger than for the NLR or PLR in literature meta-analysis, may indicate that integrating neutrophils, platelets, and lymphocytes into one index (SII) provided a stronger prognostic signal. This finding supports the results of Sun et al. (35) who reported SII as a stronger independent predictor of survival than NLR or LMR in their multivariable modeling.

Besides SII, other composite biomarkers of nutritional-inflammatory status have emerged. Notably, the prognostic nutritional index and fibrinogen-to-albumin (36) ratio is an index that describes the connection between coagulation and nutritional status. In a cohort of 427 patients resected for GBC, a high preoperative FAR was found to be an independent predictor of poorer OS and had better predictive accuracy than NLR, PLR or other immune-response biomarkers on time-dependent ROC analysis (37).

The prognostic implications of the SII in GBC may reflect the important contribution of systemic inflammation and immune response to the pathophysiology of malignancy. Each of these factors has a mechanistic role in carcinogenesis and tumor progression. Neutrophils, as pivotal effector cells of the innate immune system, can promote cancer biology in the tumor microenvironment (36). The neutrophils release inflammatory cytokines such as interleukin-6 and tumor necrosis factor-alpha and proteases that might contribute to genetic instability, angiogenesis, and tumor cell proliferation (38). Also, tumor-associated neutrophils can suppress T cell functions and promote pre-metastatic niche formation (39). Platelets have a multi-faceted role in the dissemination of cancer as well. The platelets protect the circulating tumor cells from immune destruction and can release a large amount of various growth factors, including vascular endothelial growth factor, platelet-derived growth factor, and transforming growth factor-beta, that promote angiogenesis, tumor development, epithelial-mesenchymal transition and metastasis (40). Thrombocytosis is associated with advanced disease and poorer outcomes across diverse malignancies, which may reflect platelets’ pro-tumorigenic activity within the circulation and microvasculature. (12, 41, 42).

In contrast, lymphocytes are crucial for the anti-tumor immunity and tumor-infiltrating lymphocytes can directly kill tumor cells, and their presence typically suggests a more favorable prognosis (43, 44). Lymphopenia indicates a compromised capacity for immune surveillance by the host (45). Low lymphocyte counts in patients with GBC are associated with impaired anti-tumor immunosurveillance and may contribute to more aggressive tumor behavior and earlier recurrence (46). Therefore, SII index that rises with neutrophils and platelets but falls with lymphocytes reflects aggressive cancer behavior.

Our findings in GBC consist with the oncology literature, in which the SII has emerged as a prognostic biomarker across diverse malignancies. The index was first proposed in hepatocellular carcinoma (47) and validated as a prognostic biomarker in cancers of the digestive tract, genitourinary system, lung cancer, and others (15, 48, 49). In gastric cancer, Qiu et al. (50) conducted a meta-analysis of eight studies (n=4,236) and found that high preoperative SII predicted poorer OS (HR 1.40; 95% CI 1.08–1.81) and was associated with advanced TNM stage and lymph node metastasis. Despite every tumor type does have individual characteristics, a common feature across malignancies is that a high SII indicates a less favorable tumor microenvironment and a more aggressive clinical course.

The level of between-study heterogeneity was moderate, and some design and method factors may contribute. First, SII cut-offs varied widely and were, which create threshold effects that affect both who is classified as “high SII “ and the magnitude of the HR. Second, differences in case-mix such as the proportion of stage III–IV disease and lymph-node positivity may change hazard estimates. Third, preoperative inflammatory status may be influenced by biliary infection or obstruction, cholangitis, biliary drainage, and other comorbidities, and the lack of adjustment for these factors in some studies likely resulted in residual confounding. Although funnel plot inspection and Egger’s test did not suggest small-study effects, the limited number of studies reduces power to detect asymmetry. Together, these factors explain the heterogeneity, observed in the studies, despite the consistency of the overall effect direction.

It is important to recognize potential limitations of current study. First, the evidence base comprises retrospective cohort studies, which may lead to selection bias and residual confounding despite adjustment. Second, most studies used SII cut-offs derived from data without external validation, which may lead to overfitting and limit applicability. Third, all cohorts were conducted in a single country (China) over a long accrual period, during which surgical techniques and adjuvant therapies evolved, potentially limiting generalizability to other settings and contemporary practice. Fourth, measurement variability, including the timing of preoperative blood sampling, measurement differences between laboratories, and intercurrent inflammation may have introduced misclassification. Although publication bias was not detected, the small number of studies warrants cautious interpretation.

The demonstration that a high preoperative SII predicts poor survival may have important and practical implications for GBC management. SII is obtained from routine blood counts and can be reported before any treatment intervention. Therefore, SII is a helpful tool for risk stratification without additional burden on the patient or healthcare system. Clinicians could use the SII to identify high-risk patients and counsel them about their elevated risk of recurrence or death despite surgical resection. Such a patient may be considered for more aggressive therapy or closer surveillance. We suggest that integrating the SII into prognostic models may improve predictive accuracy. Li et al. (30) proposed a nomogram that incorporated SII, tumor stage, differentiation, CA19-9, and margin status, and the nomogram was a better model than the traditional staging system for stratifying patients’ outcomes.

Another potential implication involves preoperative optimization. Given that SII reflects systemic inflammation and immune status, modification of these pathways may be explored as adjunctive therapies for surgery, and those with high SII may benefit from optimization of immuno-nutrition, anti-inflammatory regimens, and neoadjuvant therapies. However, because these observations are hypothesis-generating and require prospective validation, clinicians should consider measuring preoperative SII in patients with resectable GBC to inform risk stratification.

## Conclusion

This systematic review and meta-analysis suggests that a high preoperative SII may be an adverse prognostic factor in resectable GBC, associated with poor survival outcome. SII is a low-cost and readily available biomarker that can aid in risk stratification and reflects a state of severe inflammation associated with immunosuppression that facilitates cancer progression. Although there is moderate heterogeneity across studies due to differences in thresholds, the overall evidence is consistent.

## Abbreviations

GBC: gallbladder cancer
SII: systemic immune–inflammation index
OS: overall survival
HR: hazard ratio
CI: confidence interval
NLR: neutrophil-to-lymphocyte ratio
PLR: platelet-to-lymphocyte ratio
MLR: monocyte-to-lymphocyte ratio
FAR: fibrinogen-to-albumin ratio
TNM: tumor–node–metastasis
NOS: Newcastle–Ottawa Scale
PRISMA: Preferred Reporting Items for Systematic Reviews and Meta-Analyses

## Declarations Ethics approval

This study is a systematic review and meta-analysis of previously published studies and did not involve the collection of new data from human participants or animals. According to institutional and national regulations, ethics approval was not required.

## Human Ethics and Consent to Participate declarations

Not applicable. This study is a systematic review and meta-analysis of previously published studies and did not involve the collection of new data from human participants or animals. According to institutional and national regulations, ethics approval and consent to participate were not required.

## Consent for publication

Not applicable. This article does not contain any individual person’s data in any form (including images, videos, or case details) that would require consent for publication.

## Authors’ contributions

M.S and M.B. conceived and designed the study. M.S, A.A.G. and S.M.M.M. performed the literature search and study selection. M.S.S. and A.V. extracted the data and performed the statistical analyses. M.S drafted the manuscript. M.B. and A.V. critically revised the manuscript for important intellectual content. All authors interpreted the data, read and approved the final manuscript, and agree to be accountable for all aspects of the work.

## Data Availability

All data produced in the present study are available upon reasonable request to the authors.

All data generated or analyzed during this study are included in this published article and its supplementary information files. The datasets used and/or analysed during the current study are available from the corresponding authors on reasonable request.

## Acknowledgements

Not applicable.

## Authors’ information

Not applicable

## Clinical trial number

Not applicable.

