## Supplementary for "Prognostic value of the preoperative systemic immune-inflammation index for overall survival after Surgical resection in gallbladder cancer: a systematic review and meta-analysis"

Table S1: Search Strategy

| Database | Search Strategy |
|---|---|
| **PubMed** | (("Systemic Immune-Inflammation" OR "Systemic Immune Inflammation" OR "SII") OR ("platelet"[Title/Abstract] AND "neutrophil"[Title/Abstract] AND "lymphocyte"[Title/Abstract])) AND (("Gallbladder Neoplasms"[Mesh]) OR ("gallbladder cancer"[Title/Abstract] OR "gall bladder cancer"[Title/Abstract] OR "gallbladder carcinoma"[Title/Abstract] OR "gall bladder carcinoma"[Title/Abstract] OR "gallbladder neoplasm"[Title/Abstract] OR "gall bladder neoplasm"[Title/Abstract] OR "gallbladder adenocarcinoma"[Title/Abstract] OR "gallbladder squamous cell carcinoma"[Title/Abstract] OR "gallbladder adenosquamous carcinoma"[Title/Abstract] OR "gallbladder mucinous carcinoma"[Title/Abstract] OR "gallbladder signet ring carcinoma"[Title/Abstract])) |
| **Embase** | ('systemic immune-inflammation' OR 'systemic immune inflammation'/exp OR 'systemic immune inflammation' OR 'tuberculin'/exp OR 'tuberculin' OR ('platelet':ti,ab,kw AND 'neutrophil':ti,ab,kw AND 'lymphocyte':ti,ab,kw)) AND ('gallbladder cancer':ti,ab,kw OR 'gall bladder cancer':ti,ab,kw OR 'gallbladder carcinoma':ti,ab,kw OR 'gall bladder carcinoma':ti,ab,kw OR 'gallbladder neoplasm':ti,ab,kw OR 'gall bladder neoplasm':ti,ab,kw OR 'gallbladder adenocarcinoma':ti,ab,kw OR 'gallbladder squamous cell carcinoma':ti,ab,kw OR 'gallbladder adenosquamous carcinoma':ti,ab,kw OR 'gallbladder mucinous carcinoma':ti,ab,kw OR 'gallbladder signet ring carcinoma':ti,ab,kw) |
| **Scopus** | ( ALL ( "Systemic Immune-Inflammation" OR "Systemic Immune Inflammation" OR "SII" ) OR TITLE-ABS-KEY ( "platelet" AND "neutrophil" AND "lymphocyte" ) AND TITLE-ABS-KEY ( "gallbladder cancer" OR "gall bladder cancer" OR "gallbladder carcinoma" OR "gall bladder carcinoma" OR "gallbladder neoplasm" OR "gall bladder neoplasm" OR "gallbladder adenocarcinoma" OR "gallbladder squamous cell carcinoma" OR "gallbladder adenosquamous carcinoma" OR "gallbladder mucinous carcinoma" OR "gallbladder signet ring carcinoma" ) ) |
| **Web of Science** | "Systemic Immune-Inflammation" OR "Systemic Immune Inflammation" OR "SII" OR ("platelet" AND "neutrophil" AND "lymphocyte") (Topic) AND "gallbladder cancer" OR "gall bladder cancer" OR "gallbladder carcinoma" OR "gall bladder carcinoma" OR "gallbladder neoplasm" OR "gall bladder neoplasm" OR "gallbladder adenocarcinoma" OR "gallbladder squamous cell carcinoma" OR "gallbladder adenosquamous carcinoma" OR "gallbladder mucinous carcinoma" OR "gallbladder signet ring carcinoma" (Topic) |

Table S2. Risk of Bias Assessment using NOS

| Study (year) | Design | Selection (0-4★) | Comparability (0-2★) | Outcome/Exposure (0-3★) | Overall Score (0-9★) | Quality (High/Moderate/Low) |
|---|---|---|---|---|---|---|
| Huang (2025)[20] | Retrospective Cohort | 4/4★ | 2/2★ | 1/3★ | 7/9★ | High |
| Li (2023)[11] | Retrospective Cohort | 4/4★ | 2/2★ | 2/3★ | 8/9★ | High |
| Liu (2025)[21] | Retrospective Cohort | 4/4★ | 2/2★ | 3/3★ | 9/9★ | High |
| Sun (2020)[29] | Retrospective Cohort | 4/4★ | 2/2★ | 2/3★ | 8/9★ | High |
| Yin (2024)[22] | Retrospective Cohort | 4/4★ | 2/2★ | 1/3★ | 7/9★ | High |
| Li (2021)[28] | Retrospective Cohort | 4/4★ | 2/2★ | 2/3★ | 8/9★ | High |
| Chen (2021)[23] | Retrospective Cohort | 4/4★ | 2/2★ | 1/3★ | 7/9★ | High |

Table S3: GRADE Summary of Findings for the association between high preoperative SII and overall survival after Surgical surgery for gallbladder cancer

| Domain | Judgment | Rationale | Decision |
|---|---|---|---|
| **Risk of bias** | **Serious** | Retrospective cohorts; variable adjustment for confounders (e.g., TNM, LN status, CA19-9); several studies used data-driven cut-offs (ROC/X-tile); one HR reconstructed from KM/log-rank. | **Downgrade 1** |
| **Inconsistency** | Not serious | Direction consistent across studies; heterogeneity modest ($I^2 \approx 26\%$); sensitivity analyses stable. | No downgrade |

| | | | |
|---|---|---|---|
| **Indirectness** | Not serious | Population, exposure (preop SII), and outcome (OS) directly match the review question. | No downgrade |
| **Imprecision** | Not serious | Pooled HR **2.17** with 95% CI **1.55–2.79** excludes no effect and does not approach trivial effects. | No downgrade |
| **Publication bias** | Not detected | Funnel/Egger do not indicate small-study effects (power limited but no clear asymmetry). | No downgrade |
| **Large effect (upgrade)** | Not applied | Although HR >2, residual confounding and data-driven thresholds argue against upgrading. | — |
| **Dose–response (upgrade)** | Not assessed | Continuous-SII analyses suggest monotonic trends in some studies, but not uniformly reportable. | — |

Summary of effect: HR 2.17 (95% CI 1.55–2.79; 7 studies; 2,153 participants)

Overall certainty: Moderate

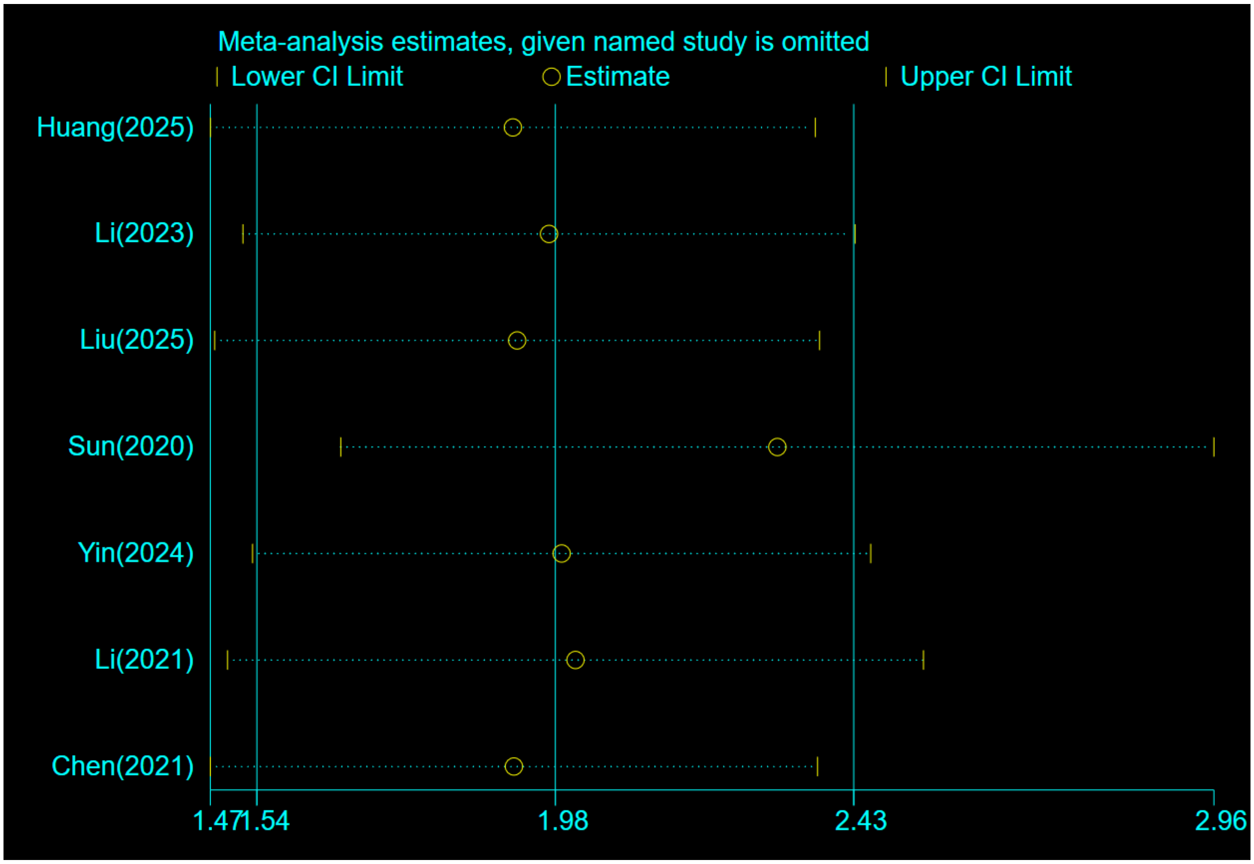


Figure S1: Leave-one-out sensitivity analysis showing the pooled HR after omitting each study in turn.
